# Endocannabinoid response in acute ischemic stroke: elevated 2-arachidonoylglycerol

**DOI:** 10.1101/2020.10.29.20222646

**Authors:** Marina Buciuc, Gian Marco Conte, Eugene L. Scharf

**Affiliations:** Departments of Neurology, Mayo Clinic, Rochester, MN, USA; Departments of Neurology Radiology, Mayo Clinic, Rochester, MN, USA

**Keywords:** Acute ischemic stroke, Transient ischemic attack, Endocannabinoids, Cell signaling, Lipids

## Abstract

**Background and purpose:** Endocannabinoids are hypothesized to have anti-inflammatory and neuroprotective properties and hold therapeutic potential in the acute phase response mechanisms during acute cerebral ischemia and closed head injury. We set to describe the plasma levels of endocannabinoids and related ethanolamides during acute and subacute phases of cerebral ischemia.

**Methods:** We conducted a prospective observational study of plasma endocannabinoid levels in patients with acute ischemic stroke or transient ischemic attack. Two blood samples were collected: T1 (<12 hours from symptom onset) and T2 (>24 hours from symptom onset). N-arachidonoylethanolamine (AEA), 2-arachidonoylglycerol (2-AG), palmitoylethanolamide (PEA) and oleoylethanolamide (OEA) were quantified by liquid chromatography mass spectrometry.

**Results:** Twenty-three patients met inclusion criteria. Median (interquartile range): Age – 76 years (60-81); body mass index - 25.6 (23.6-30.4); National Institutes of Health Stroke Scale score-5(3-13); infarct volume - 1.4 cm^3^ (0.5-8.6). Higher 2-AG levels at T1 were correlated with smaller infarct volumes (Spearman ƿ=-0.48, *p*=0.0206). Levels of 2-AG were elevated at T2 compared to T1 in 48% of patients (median difference - 310.3nM, 95% CI 194.1-497.3; *p*=0.001); AEA, PEA and OEA did not differ between T1 and T2, *p*>0.05. Patients with elevated 2-AG at T2 had larger infarct volumes, *p*=0.0178, lower frequency of embolectomy performed, *p*=0.0373, but no difference in neurological disability 90 days after the ischemic event compared to patients without 2-AG elevation.

**Conclusion:** 2-AG increases significantly in early phases of ischemic stroke. The final mechanistic role of 2-AG in acute ischemic stroke is to be determined in further studies.

## INTRODUCTION

Cerebrovascular disease, stroke being the most common, represents one of the leading causes of death in adults worldwide regardless of country income and attributes to almost 10% of all deaths^1^. Stroke survivors as well as patients who experienced transient ischemic attack (TIA) are at a higher risk of suffering from the recurrence of cerebrovascular event which can result in death, accumulating disability limiting their independence in the activities of daily living as well as experiencing financial hardships related to hospital costs and care home institutionalization as early as 5 years after the event^2, 3^. Therefore, the need for improvement in acute treatment, discovery of novel agents aimed for better neurological recovery and secondary prevention in stroke and TIA patients is paramount.

Recently, the human endocannabinoid (eCB) system has gained interest as a therapeutic target for ischemic stroke given its pleiotropic role in physiology and abundance of eCB receptors in central nervous system^4^. The human eCB system is a distributed network of cellular membrane-derived bioactive lipid amides believed to provide homeostasis to vascular tone, immune system, hemostasis, metabolic balance, and neurotransmission via widespread actions through G-coupled protein, nuclear, and ligand-gated ion channel receptors ^5^. The main components of the eCB system are the derivatives of acyl phosphatidylcholine – the N-acyl ethanolamines, most notably arachidonoyl ethanolamide, or AEA; and secondly, the derivatives of inositol phospholipids – the diacylglycerols, most notably the 2-arachidonoylglycerol (2-AG). The role of eCB signaling in acute ischemic stroke is limited to a case report ^6^ and a single series^7^ suggesting early elevations in AEA and related N-palmytoilethanolamide (PEA) and their possible correlation with the severity of stroke. Preclinical studies have reported potential involvement of AEA and related N-oleoylethanolamide (OEA) in preservation of blood-brain barrier during acute cerebral ischemia^8^, reduction of inflammation and promotion of neurogenesis after transient focal ischemic event^9, 10^. Neuroprotective effects of increased 2-AG levels in experimental brain and spinal cord injury^11, 12^ via possible effect on glial cells^13^ and/or inflammation reduction.

Given the neuroprotective potential suggested by eCB lipids further characterization is warranted. The following exploratory study sought to describe signaling of eCB and related ethanolamides in acute to subacute cerebral ischemic events over time in a clinical cohort.

## METHODS

### Study design and participants

We conducted a prospective longitudinal observational study using patients who presented with acute ischemic stroke or TIA symptoms to the Mayo Clinic Hospital (Saint Mary’s campus) in Rochester, MN or referring health system emergency room. All patients were screened and enrolled between November 1^st^, 2018 and December 1^st^, 2019. We identified patients who met the following inclusion criteria: 1) presented to the hospital within twelve hours of symptom onset or last known well (LKW); 2) had National Institutes of Health Stroke Scale (NIHSS)^14^ score ≥2 or NIHSS score = 0 and ABCD2 score ≥4^15, 16^ for suspected TIA on admission; 3) initial non-contrast CT head excluded the presence of primary intracerebral hemorrhage or subarachnoid hemorrhage. Patients who were <18 years old, had fever >38.0°C and/or white blood cell count> 16,000cells/µl, had a history or suspicion of active endocarditis, active malignancy, inflammatory vasculopathy (e.g. Moya-Moya, Susac’s syndrome), systemic inflammatory disease, connective tissue disease, hypercoagulable state, seizure at presentation, had recent consumption or inhalation of exogenous cannabinoids, or pregnancy were excluded.

Out of 253 screened patients who presented with acute stroke or TIA symptoms, 226 were excluded: LKW>12 hours ago, n=56; not enough evidence for stoke or TIA on initial neurological exam, n=27; initial non-contrast CT head showed intracerebral or subarachnoid hemorrhage, n = 14; met one or more exclusion criteria, n=123; unable to consent due to language barrier, n = 6. Twenty-seven patients met our inclusion and exclusion criteria and were consented. On admission patients underwent emergency neurological evaluation according to accepted practice standards^17, 18^. Dynamics of eCB signaling were investigated in pooled venous blood by phlebotomy. The first blood draw (T1) was performed within 12 hours of LKW and intended to measure eCB signaling during acute ischemic stroke. The second time point (T2) for blood sample collection was chosen closest to discharge time of the patients in order to allow for return to the closest to the baseline state, was at least 24 hours after the symptom onset or LKW and no longer than 7 days after and was intended to measure eCB signaling during subacute phase of cerebral ischemic event. Demographic and clinical characteristics as well as relevant laboratory markers were abstracted from the medical record (MB). Screening logs were kept.

Mechanism of acute ischemic event and modified Rankin scale (mRS) score^19, 20^ for neurological disability 90 days after the event have been determined by retrospective chart review conducted by a blinded to imaging and laboratory data board certified neurologist (ELS).

### Standard protocol approvals, registrations and patient consents

This study has been approved by the Mayo Clinic institutional review board, and all participants and/or their proxies signed a written informed consent form before taking part in any research activities in accordance with the Declaration of Helsinki. Due to time-sensitivity of the sample acquisition a deferred consent was approved for the present study by the Mayo Clinic institutional review board which allowed collection a blood sample from a qualifying candidate when a written informed consent was not possible to obtain within 12 hour of LKW. In those cases written informed consent was still required from the participants and/or their proxies after the sample collection before any data abstraction and/or analyses were performed; otherwise, the collected samples were destroyed.

### Biochemical Analysis

Three milliliters of venous blood were collected by phlebotomy into chilled EDTA-containing tubes, centrifuged for plasma separation, aliquoted in 1.5 ml tubes, snap-frozen and immediately stored at -80 C° until further analyses^21^. The levels of the following endocannabinoids and naturally occurring ethanolamide lipids were assessed: 2-arachidonoylglycerol (2-AG); N-arachidonoylethanolamide (AEA); N-oleoylethanolamide (OEA); N-palmytoilethanolamide (PEA). Quantitative assays were performed by liquid chromatography mass spectrometry (LC/MS) as previously described with a few modifications^22-24^. Briefly, 50µl of internal standard solution containing d4-OEA, d4-AEA, d4-PEA, and d5-2-AG was added to 200µl of plasma. Proteins were removed by adding 1ml of 50:50 heptane:ethyl acetate solution to the sample mixture. The mixture was vortexed for 10 minutes prior to being centrifuged for 10 minutes. The organic layer was then transferred to a separate vial and dried under speedvac. After drying the samples were re-suspended in 30µL of methanol before being analyzed on a Nexera X2 UPLC module (Shimadzu Scientific Instruments, Columbia, MA) coupled with a SCIEX 6500 triple quadrupole mass spectrometer (SCIEX, Framingham, MA). Metabolites were separated on an Acquity UPLC BEH C18, 1.7 μm, 2.1 × 150 mm column (Waters Corp, Milford, MA), held at 25 °C, using 0.1% acetic acid as mobile phase A, and 9:1 mixture acetonitrile to isopropyl alcohol as mobile phase B. A 16 minute runtime with a flow rate of 0.3 mL/min and a gradient elution (30% to 90% mobile phase A) was utilized to separate the analytes. The mass spectrometer was operated in positive electrospray mode, monitoring mass transitions of *m/z* 348>62 for AEA, 326>62 for OEA, 300>62 for PEA, and 379>287 for 2-AG. Concentrations of all analytes were measured against a 10-point calibration curve.

### Neuroimaging Analysis

For each patient, manual segmentations of the infarct areas were obtained using the software RIL-Contour^25^. Areas of restricted diffusion were segmented on DWI images without using a priori thresholds. To confirm true restricted diffusion and avoid the T2 shine through phenomena^26^, apparent diffusion coefficient (ADC) maps were used as a reference to look for a corresponding area of low signal. The final segmentation included only areas that presented as hyperintense on DWI and hypointense on ADC maps. In cases where MR imaging was not performed (n=6), infarct areas were obtained using non-contrast-enhanced CT scans.

### Statistical Analysis

JMP Pro 14 statistical software package was used for data analysis (https://www.jmp.com/en_us/software/predictive-analytics-software.html). Due to the actual sample size <25 and not normally distributed data nonparametric methods were implemented for statistical analyses. The Wilcoxon rank sum test was used to compare continuous variables; while Chi-square or Fisher’s exact tests were used for categorical variables such as sex. The primary endpoint was difference in median (95% confidence interval) levels of eCBs and related ethanolamides between T1 to T2 ; the estimates and 95% confidence intervals were calculated using smoothed empirical likelihood methods for quantiles^27^. Statistical significance of difference in levels of paired measurements was assessed by two-sided Wilcoxon signed rank test with predetermined level of significance defined as any α error of <0.05. Five-fold or more change in levels of circulating plasma eCBs or ethanolamides from T1 to T2 was arbitrarily determined to be the minimum change necessary to be considered a physiological relevant event based on preclinical literature where five-fold increase in exogenously administered 2-AG resulted in significant neurological recovery from induced closed head injury compared to previous dose or vehicle^11^. We also conducted multivariate Spearman’s rho correlation analysis to look for associations between the variables. Figures were generated in R software version 3.4.2^28^. No formal power calculation was completed for enrollment, it was estimated that approximately 25 patients would provide adequate measures of central tendency for powering larger studies.

### Data availability policy

Anonymized data are available from the corresponding author upon request from any qualified investigator for purposes of replicating procedures and results.

## RESULTS

A total of twenty-three patients were included for the final analyses: two patients were excluded due to failure to obtain one of the two required blood samples; two patients were excluded due to absence of signs of justifiable cerebral ischemia during the diagnostic work-up (e.g. absence of areas of restricted diffusion on diffusion-weighted imaging (DWI) MRI scan, substantial stenosis of carotid arteries). Demographic, clinical and laboratory characteristics of the study participants are summarized in **Table 1**. The median circulating plasma levels of 2-AG were significantly higher at T2 compared to T1 (median difference - 172.5nM, 95% CI 62.3 – 306.5; *p*<0.0001); levels of AEA, OEA and PEA did not differ between the two time points, *p*<0.05 (**Figure 1, Table 2A**). We observed that eleven patients (48%) in our study had at least 5X increase in 2-AG levels from T1 to T2 and defined them as ‘reactors’ whereas the remaining thirteen patients had a modest (<5X) elevation of 2-AG, therefore defined as ‘non-reactors’. We stratified our analyses by the reactor status to evaluate whether levels of AEA, PEA and OEA varied between the two groups. Only 2-AG elevation in the reactor group remained statistically significant (*p*=0.001) whereas the levels of AEA, PEA, OEA and less than five-fold increase in 2-AG did not differ between acute and subacute stages of cerebral ischemia (**Figure 2, Table 2B, 2C**). Comparison of demographic, clinical and laboratory characteristics between reactors and non-reactors revealed larger infarct volumes in reactors (median infarct volume [interquartile range], cm^3^ – 8.6 [0.9 – 20.5] vs 0.6 [0.3 – 4.2]; *p*=0.0178) compared to non-reactors. Non-reactors had a higher frequency of embolectomy performed compared to reactors (42% [5/12] vs. 0% [0/11]; *p*=0.0373) (**Table 1**). Higher 2-AG levels at time point 1 were moderately correlated with smaller infarct volumes (Spearman ƿ = -0.48, *p*=0.0206); levels of AEA, PEA, and OEA at time point 1 were highly correlated with each other (Spearman ƿ = 0.78 [AEA and OEA], ƿ =0.85 [AEA and PEA], ƿ = 0.90 [PEA and OEA], *p*<0.0001); no other relevant and statistically significant correlations were found, *p*>0.05.

**Table 1.** Demographic, clinical and laboratory characteristics of the study participants

|  | N(%) or median (interquartile range) |  |  |  |
| --- | --- | --- | --- | --- |
|  | All<br>(n=23) | Reactors<br>(n=11)<sup>†</sup> | Non-reactors<br>(n=12) | *p*-value<sup>‡</sup> |
| **Demographic characteristics** |  |  |  |  |
| Female | 12 (52%) | 4 (36%) | 8 (67%) | 0.2203 |
| Age, years | 76 (60 – 81) | 76 (60 – 81) | 77 (57 – 87) | 0.8052 |
| BMI, kg/m<sup>2</sup> | 25.6 (23.6 – 30.4) | 28.9 (22.4 – 36.4) | 24.7 (23.7– 28.9) | 0.3401 |
| **Risk Factors** |  |  |  |  |
| Hypertension | 20 (87%) | 10 (91%) | 10 (83%) | 1.0000 |
| Diabetes mellitus | 7 (30%) | 4 (36%) | 3 (25%) | 0.6668 |
| Hyperlipidemia | 19 (83%) | 10 (91%) | 9 (75%) | 0.5901 |
| Obstructive sleep apnea | 6 (26%) | 4 (36%) | 2 (17%) | 0.3707 |
| Cardiac disease | 15 (65%) | 8 (73%) | 7 (58%) | 0.6668 |
| Prior stroke | 8 (35%) | 4 (36%) | 4 (33%) | 1.0000 |
| Deep vein thrombosis | 3 (13%) | 2 (18%) | 1 (8%) | 0.5901 |
| **Medications prior to admission** |  |  |  |  |
| Antiplatelet therapy | 13 (57%) | 8 (73%) | 5 (42%) | 0.2138 |
| Anticoagulation therapy | 4 (17%) | 1 (9%) | 3 (25%) | 0.5901 |
| Statin therapy | 10 (44%) | 6 (55%) | 4 (33%) | 0.4136 |
| **Acute ischemic event characteristics** |  |  |  |  |
| NIHSS score | 5 (3 – 13) | 5 (4 – 8) | 4 (2 – 22) | 1.0000 |
| Infarct volume, cm<sup>3</sup> | 1.4 (0.5 – 8.6) | 8.6 (0.9 – 20.5) | 0.6 (0.3 – 4.2) | 0.0178 |
| Systolic BP, mmHg | 158 (137– 167) | 160 (149 – 173) | 146 (135 – 166) | 0.1567 |
| Diastolic BP, mmHg | 89 (75 – 99) | 89 (75 – 96) | 86 (71 – 107) | 0.8535 |
| IV-tPA administered | 10 (44%) | 5 (46%) | 5 (42%) | 1.0000 |
| Embolectomy performed | 5 (22%) | 0 (0%) | 5 (42%) | 0.0373 |
| Modified Rankin scale | 2 (0 – 3) | 2 (0 – 3) | 1.5 (0 – 3) | 0.8742 |
| **Mechanism of the acute ischemic event** |  |  |  |  |
| Cardioembolic | 8 (35%) | 3 (27%) | 5 (42%) | 0.6668 |
| Large vessel occlusion | 4 (17%) | 2 (18%) | 2 (17%) | 1.0000 |
| ESUS | 7 (31%) | 4 (37%) | 3 (25%) | 0.6668 |
| Other<sup>§</sup> | 4 (17%) | 2 (18%) | 2 (17%) | 1.0000 |
| **Laboratory results** |  |  |  |  |
| Total cholesterol, mg/dL | 161 (129 – 184) | 169 (146 – 204) | 146 (125 – 181) | 0.1997 |
| LDL, mg/dL | 79 (67 – 122) | 104 (75 – 124) | 75 (57 – 121) | 0.2493 |
| HDL, mg/dL | 45 (34 – 56) | 47 (30 – 55) | 43 (35 – 58) | 0.9214 |
| Triglycerides, mg/dL | 114 (75 – 152) | 143 (70 – 161) | 108 (77 – 134) | 0.4116 |
| Hemoglobin A1c, % | 5.7 (5.4 – 6.2) | 5.7 (5.4 – 6.0) | 5.7 (5.2 – 6.4) | 1.0000 |
| Ejection fraction, % | 60 (51 – 65) | 55 (43 – 60) | 65 (53 – 69) | 0.0927 |
| **Time from last known well to blood sample acquisition** |  |  |  |  |
| T1, hours | 8.5 ( 6.8 – 11.0) | 8.8 (6.0 – 11.0) | 8.0 (6.8 – 9.8) | 0.6647 |
| T2, hours | 45 (33 – 56) | 42 (29 – 79) | 49 (39 – 54) | 0.6665 |

Abbreviations: BMI, body mass index; NIHSS, National Institutes of Health Stroke Scale; IV-tPA , intravenous tissue plasminogen activator; ESUS, embolic stroke of undetermined source
<sup>†</sup>Reactors – participants with 5X or more increase in levels of 2-arachidonoylglycerol between T1 and T2
<sup>‡</sup>For categorical variables, *p*-values are from Fisher's Exact Test; for continuous - from Wilcoxon rank sum test
<sup>§</sup>Other mechanisms: dissection (n=1), hypotension (n=1), transient ischemic attack (n=1), small vessel occlusion (n=1)

**Table 2.** Median difference in levels of circulating endocannabinoids between the two time points

| Endocannabinoids | T1<br>( n =24)<sup>†</sup> | T2<br>( n =24) | Median difference<br>(95% CI)<sup>‡</sup> | p-value<sup>§</sup> |
| --- | --- | --- | --- | --- |
| **A. All study participants (n=23)** |  |  |  |  |
| 2-AG, nMol | 9.2 (5.1 – 18.5) | 235.6 (16.9 – 412) | 172.5 (62.3, 306.5) | <0.0001 |
| AEA, nMol | 0.9 (0.7 – 1.3) | 0.8 (0.5 – 1.1) | -0.13 (-0.44 , 0.05) | 0.1036 |
| OEA, nMol | 4.9 (3.1 – 8.5) | 4.4 (2.8 – 5.8) | -0.16 (-2.11, 1.06) | 0.4776 |
| PEA, nMol | 8.3 (6.8 – 10.1) | 7.7 (4.8 – 8.6) | -0.46 (-2.20, 0.75) | 0.2376 |
| **B. Study participants with 5X and more increase in 2-AG levels from T1 to T2 (n=11)** |  |  |  |  |
| 2-AG, nMol | 8.0 (5.2 – 16.9) | 308.5 (176.4 – 540.1) | 310.3 (194.1, 497.3) | 0.0010 |
| AEA, nMol | 1.2 (0.8 – 1.3) | 0.9 (0.6 – 1.4) | -0.12 (-0.53 , 0.08) | 0.3594 |
| OEA, nMol | 4.9 (2.5 – 8.5) | 4.8 (3.1 – 5.8) | 0.40 (-1.87, 1.53) | 0.9219 |
| PEA, nMol | 8.7 (7.0 – 10.1) | 8.0 (5.8 – 9.3) | 0.01 (-1.92, 1.27) | 0.7520 |
| **C. Study participants with less than 5X increase in 2-AG levels from T1 to T2 (n=12)** |  |  |  |  |
| 2-AG, nMol | 10.9 (5.0 – 141.3) | 19.3 (5.0 – 401.1) | 30.4 (-19.3, 242.8) | 0.2334 |
| AEA, nMol | 0.8 (0.5 – 1.4) | 0.7 (0.4 – 0.9) | -0.21 (-0.68, 0.16) | 0.2373 |
| OEA, nMol | 5.6 (3.1 – 10.1) | 4.0 (2.7 – 6.6) | -1.16 (-4.90, 1.29) | 0.3013 |
| PEA, nMol | 7.2 (6.4 – 10.1) | 7.4 (4.5 – 8.1) | -1.28 (-3.92, 0.89) | 0.1455 |

Abbreviations: nMol, nanomolar; CI, confidence interval; 2-AG, 2-arachidonoylglycerol; AEA, N-arachidonoylethanolamide; OEA, N-oleoylethanolamide; PEA, N-palmytoilethanolamide
<sup>†</sup>Data shown for T1 and T2 are median (interquartile range)
<sup>‡</sup>Median difference and 95% confidence intervals are from smoothed empirical likelihood quantiles
<sup>§</sup>p-values shown are from Wilcoxon signed rank test, $p < 0.05$ was considered statistically significant

**Figure 1.**
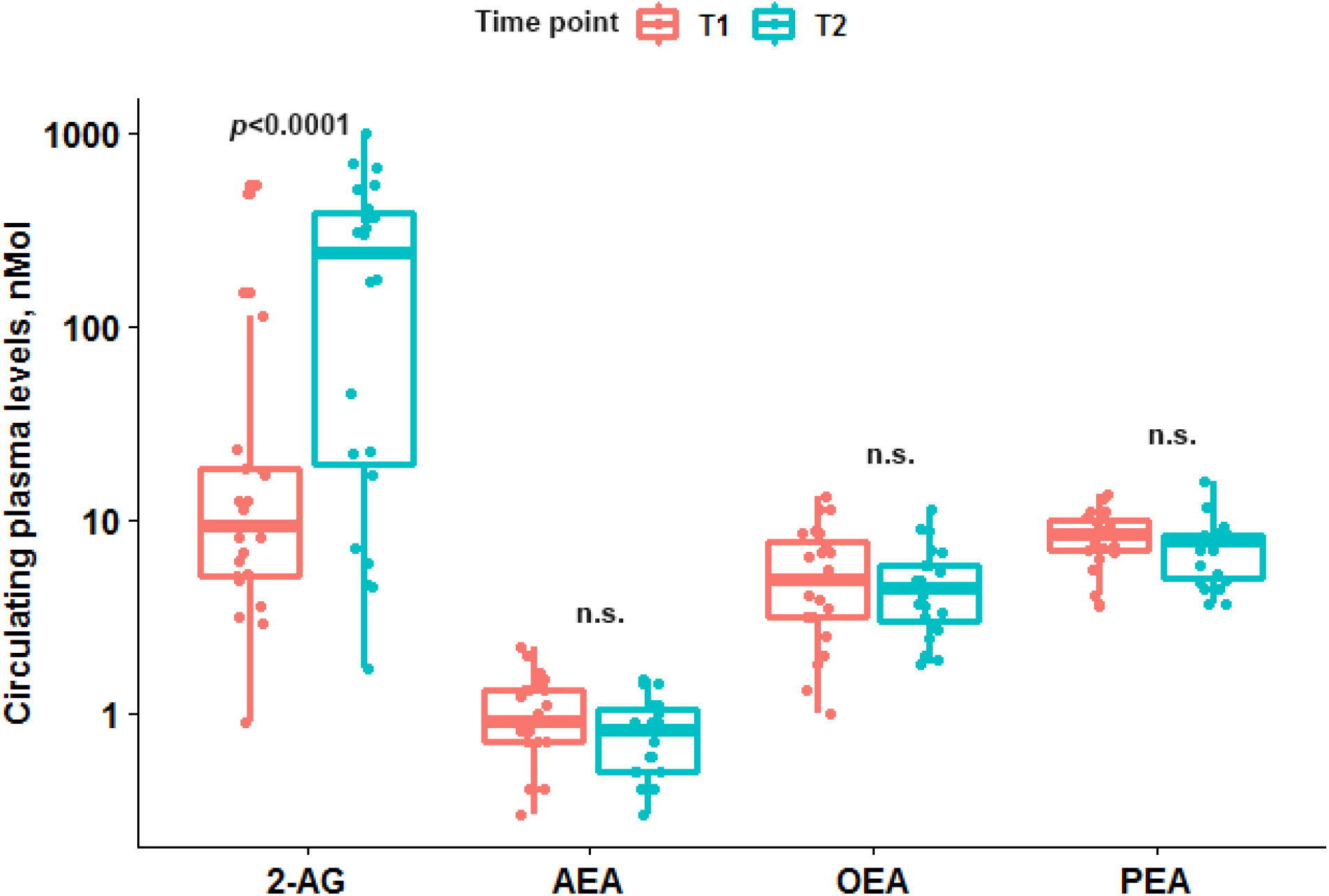
Box plots showing difference in levels of endocannabinoids and related ethanolamides between two time points. Coral box plots represent circulating plasma levels of endocannabinoids at time point 1 (<12 hours from the symptom onset) and turquoise box plots represent plasma levels at time point 2 (> 24 hours from the symptom onset). The line in box plots represents the median, interquartile range and the whiskers correspond to minimum and maximum values. Abbreviations: 2-AG – 2-arachidonoylglycerol; AEA – N-arachidonoylethanolamide (AEA); OEA – N-oleoylethanolamide; PEA – N-palmytoilethanolamide; n.s. – not significant, *p*≥0.05

**Figure 2.**
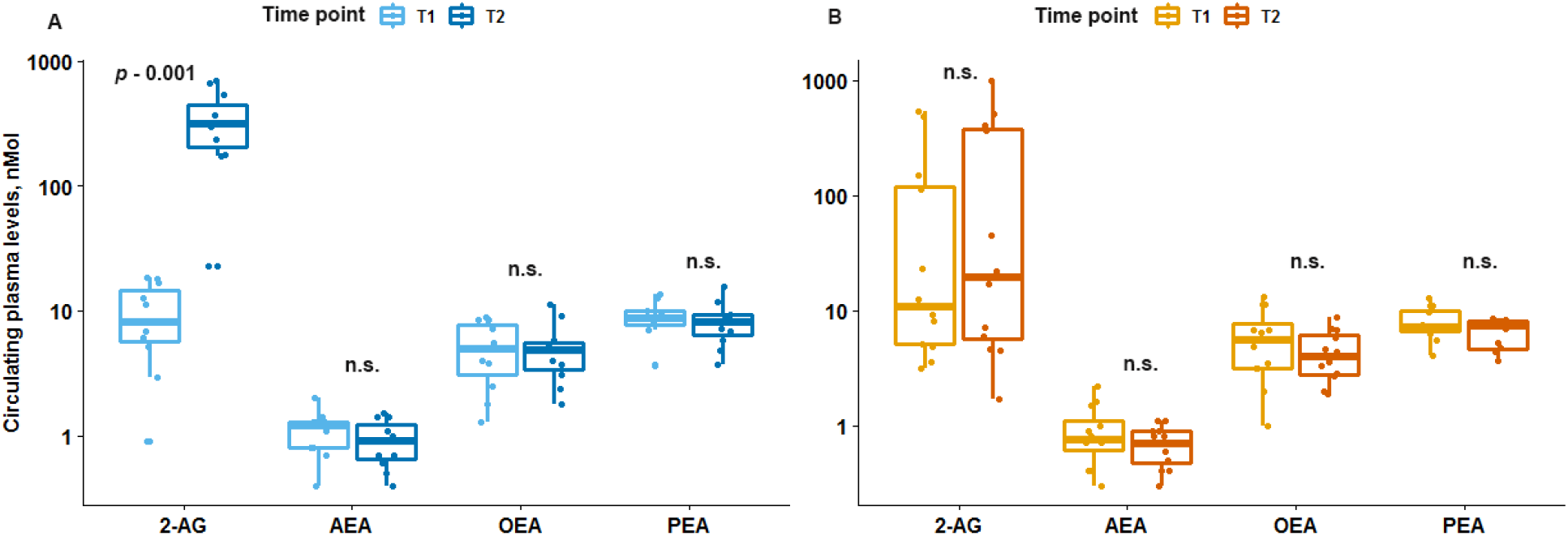
Box plots showing difference in levels of endocannabinoids and related ethanolamides between two time points for reactors and non-reactors. Reactors had a significant increase in 2-AG levels at time point 2 (panel **A**) compared to non-reactors (panel **B**). Light blue and mustard box plots represent circulating plasma levels of endocannabinoids at time point 1 (<12 hours from the symptom onset); dark blue and orange box plot represent plasma levels at time point 2 (> 24 hours from the symptom onset). The line in box plots represents the median, interquartile range and the whiskers correspond to minimum and maximum values. Abbreviations: 2-AG – 2-arachidonoylglycerol; AEA – N-arachidonoylethanolamide (AEA); OEA – N-oleoylethanolamide; PEA – N-palmytoilethanolamide; n.s. – not significant, *p*≥0.05

## DISCUSSION

Therapeutic management of acute cerebral ischemia, especially in patients who do not qualify for thrombolytic therapy and/or embolectomy, is limited and represents an unmet medical necessity. This is the first clinical study to our knowledge that demonstrates that the endocannabinoid 2-AG increase in patients with large infarcts who did not undergo embolectomy.

Our findings of elevated levels of 2-AG are consistent with preclinical murine studies which also showed an increase in 2-AG after acute brain or spinal cord injury^11, 12, 29^ including acute cerebral ischemia^30^. Moreover, we found a correlation between higher levels of 2-AG in the first hours of acute ischemic event and smaller infarct size which is in accordance with previous murine studies that reported reduction in lesion expansion and smaller infarct volumes with endogenous elevation or exogenous administration of 2-AG^12, 30^. The mechanism behind this association might be related to the role of 2-AG in inhibition of inflammation and preservation of blood-brain barrier^11, 29^, inhibition of neurotransmission as well as vasodilation^31, 32^ – all of which might aid in faster reperfusion and decrease of edema and cell injury in the ischemic brain tissue. Interestingly, when 2-AG levels were analyzed over time, we observed that not all patients had a substantial increase of 2-AG levels in the subacute vs. acute phase of the ischemic event. When we compared these two groups of patients, we noted that those with significant increase of 2-AG over time had larger infarct volumes. This observation calls into question the role of the elevation of 2-AG levels in the subacute phase of ischemic stroke: is 2-AG neuroprotective and its increase is proportional to the size of the injury or does 2-AG contribute to the thrombosis*?*

*In vitro* experiments have demonstrated that 2-AG acts as a platelet agonist, promotes platelet activation and aggregation and is also released by the activated platelets^33 34^; this finding was validated *in vivo* with a murine study where exogenous 2-AG administration (micromolar doses) induced platelet aggregation and worsened cerebral blood flow in the first hour of middle cerebral artery occlusion^35^. In our study we also observed that all patients who underwent embolectomy (all procedures resulted in successful recanalization) did not have significant elevations of 2-AG in the subacute phase. Therefore, 2-AG may serve multiple physiological roles in acute cerebral ischemia–partially attributed to its production and release by the platelets in the thrombus as well as reactive response to tissue injury in the attempt to minimize it through multiple pathways. However, the exact source of 2-AG in acute cerebral ischemia has not been determined.

We observed a trend in decrease of AEA levels between acute phase of ischemic event (<12 hours from the symptom onset) and subacute phase (≥24 hours from the symptom onset), *p*=0.1036; our finding, however, did not reach statistical significance and does not reproduce results reported in the previous studies.^6, 7^ Incongruence between our and previously reported results regarding AEA levels could be due to reduced power of detection due to a modest sample size, differences in the site of measurement (intracranial^6^ vs. peripheral), and/or differences in time points when the first blood sample was acquired. Naccarato *et al*. report significant elevated AEA levels in stroke patients when compared to control subjects only at 6 hours or less from the symptom onset and this difference disappears as early as 12 hours after LKW^7^. The median time of acquisition of first blood sample from LKW in our study was 8.5 hours; therefore, it is possible that we have missed the peak of AEA elevation in the first hours of acute ischemic event and in fact have observed the gradual decline of originally spiked AEA levels over time.

We did not observe significant changes in OEA and PEA levels between acute and subacute phases of ischemic event as reported by Schäbitz *et al*^6^ nor did we observe correlation of OEA/PEA levels in the first hours of acute ischemic event with NIHSS score^7^ or other relevant clinical parameters. Whereas eCBs and PEA have been described to have neuroprotective properties, a recent study demonstrated mutually neutralizing effects of 2-AG and PEA in presumed neuroprotection of microglia^36^. Highly correlated levels of PEA with OEA and AEA, but not 2-AG, in the acute phase of the ischemic event are also suggestive of different pathway and downstream cascade of reactions of these two neuroprotective agents. It is therefore suggestive of different, possibly mutually exclusive, mechanisms of activation of 2-AG and PEA pathway in the event of CNS injury and since in our study we observed elevation of 2-AG, it is plausible to think that PEA pathway was not activated, if not repressed, to prevent the inhibition of 2-AG effect.

Prospective design, adherence to strict inclusion/exclusion criteria as well as ample clinical and laboratory phenotyping of the participants are the strengths of this study. Our findings, however, need to be interpreted with caution due to a number of limitations. A modest sample size reduced the power of the study and our ability to detect possibly significant differences. Second, for a small number of patients infarct size has been calculated based on CT scan due to unavailability of DWI MRI which might have resulted in variability in precision; however, current literature suggests that calculations of infarct size on CT can be comparable to that on DWI MRI with evolving machine learning tools^37^. Since all the participants were Caucasian of European descent and of non-Hispanic ethnicity it reduces generalizability of our results to other ethnicities. We did not have a matched healthy control group to assess the magnitude of fluctuation of eCB levels in stroke/TIA patients compared to healthy controls. Lastly, here we report circulating plasma levels of eCBs and related ethanolamides which might or might not reflect their actual concentrations or fold change in the brain tissue.

Treatment options for acute cerebral ischemia are time-sensitive and limited to the reperfusion therapies via systemic thrombolysis and/or endovascular treatment; other mechanisms that contribute to brain tissue injury or, on the contrary, can alleviate the insult are often not targeted during therapeutic management. Neuroprotective agents capable of stabilizing or reducing infarct size are of particular interest as a novel therapeutic paradigm in the treatment of acute ischemic stroke. However, efficacious clinical studies are scarce. ^38^. Here, we provide evidence that eCB signaling, particularly 2-AG, is involved in the mechanisms of acute ischemic stroke in humans. The final mechanistic role of 2-AG involvement in stroke will need to be described in additional studies; however at present, 2-AG modulation is a rational target for neuroprotection in acute ischemic stroke.

## Acknowledgements

The authors extend their appreciation to the patients and their families who gave their time and effort to participate in the study, to Louis El Khoury, PhD, for the assistance with figure generation in R and Bradley J. Erickson, MD, PhD, for his assistance with neuroimaging analysis.

